# LONG-TERM MORPHOLOGICAL CHANGES IN THE HUMAN CORNEA LAYERS

**DOI:** 10.1101/2020.04.24.20069906

**Authors:** Merry Elizabeth Goedert, Micheline Borges Lucca, Felipe Beltrão Medeiros, Maria da Glória Silva, Renata Oliveira Soares, Gustavo Lara Rezende, Maria Regina Catai Chalita, Selma Aparecida Souza Kückelhaus

**Author notes:** Corresponding author: Selma Aparecida Souza Kuckelhaus, PhD, Faculty of Medicine, University of Brasilia, Brasilia, Federal District, Brazil, ZC 70910-000.

## Abstract

**Purpose:** To describe and compare the morphology of the corneal layers from adults and aging individuals.

**Methods:** The study was conducted with normal corneas (n = 15) of predominantly male individuals grouped into two classes (group I: < 40 years; group II: ≥ 40 years). To describe and to compare the corneas, the thickness of the layers, the integrity of the stromal lamellae and the endothelial lesions, SEM images of the central and peripheral regions of each cornea were quantified.

**Results:** The total thickness of the cornea was largest in the periphery only for group I. The Bowman, Descemet and endothelium layers of group I were thicker at the periphery of the corneas than their respective central regions, while for group II, this difference was observed only in the Bowman layer. Comparison between groups showed that the Bowman layer was thicker (central region), while the Descemet membrane and endothelium were thinner in group II; A greater number of endothelial lesions in both regions of the corneas was observed in group II, but no difference in the total number of interlamellar slits.

**Conclusions:** The morphological changes observed in group II could be related to the reduction in the pool of stem cell of limbus and, despite the corneas being from healthy individuals, these changes could compromise the success of transplantation. Our results point to an update of corneal uptake protocols and they may aid in the refractive surgeries interventions in individuals from different age groups.

## INTRODUCTION

The cornea is the main lens of the human eye, being responsible for almost three-quarters of the optical power, for the transmission and refraction of light, and it also works as a barrier to external agents (Sridhar, 2018). The normal physiology of this organ is closely related to its transparency, which depends on the structural preservation and hydration of its layers (anterior epithelium, Bowman layer, stroma, Descemet’s membrane and endothelium); functionally, it is the endothelium that acts on the active and controlled pumping of water and nutrients from the aqueous humor to the cornea matrix.

Pathologies such as keratoconus and Fuchs endothelial corneal dystrophy affect the morphofunctional structure of the cornea and consequently its transparency. Keratoconus is a corneal ectasia that arises during adolescence, progresses until the fourth decade of life and it is associated with increased levels of reactive species of oxygen, causing degradation of the stromal matrix and, thus, impairing the transparency of the cornea. It is a condition that affects 1 in 2000 individuals Arnal et al., 2011; De Bonis et al., 2011).

Fuchs’s dystrophy is more common than keratoconus, affecting 4 in 100 individuals. This pathology appears around the age of 40, causing the death of the endothelial cells (posterior corneal epithelium) and increasing the collagen production associated with Descemet’s membrane, manifesting as the corneal guttata which are irregular prominences of this layer (Elhalis et al., 2010).

Even in the absence of pathologies such as those reported above, it is observed in clinical practice that the normal aging process affects the integrity, shape and quality of the corneal layers causing edema, opacity, and even blindness (Ruberti et al., 2002). Morphofunctional changes of the cornea, whether associated to pathologies or not, require refractive surgeries or even transplants for the reestablishment of visual acuity. However, for the surgical procedures to be successful, it is essential to comprise how the corneal layers change in the aging process. Thus, considering that there are few studies applied to the detailing of the corneal architecture and the importance of the subject to orient the conducts in refractive surgeries, this study aimed to describe and compare the morphology of the corneal layers from adults and aging individuals.

## MATERIALS AND METHODS

### Individuals and ethical procedures

The study was conducted with normal corneas (n = 15) from individuals aged 49 ± 12 years, predominantly male (80%) and with different causes of death (cardiovascular, trauma, cirrhosis, hanging and electric shock). The corneas were obtained from donors of the Eye Bank of the Hospital of Base Hospital de Base of the Federal District, Brasilia, after being classified as unsuitable for transplantation and discarded. The donation was performed in accordance with the donation protocol of the Ministry of Health (DC No. 67 of September 30, 2008). The inclusion criteria in this study were corneas obtained from individuals without history of ocular diseases, while the corneas from individuals with a history of corneal neoplasia and/or with tissue trauma that could affect the morphological study were excluded. Thus, the individuals were grouped into two classes of age: group I < 40 years (n = 5; 35 ± 5 years) and group II ≥ 40 years (n = 10; 57 ± 6 years) (Table 1).

**Table 1.**
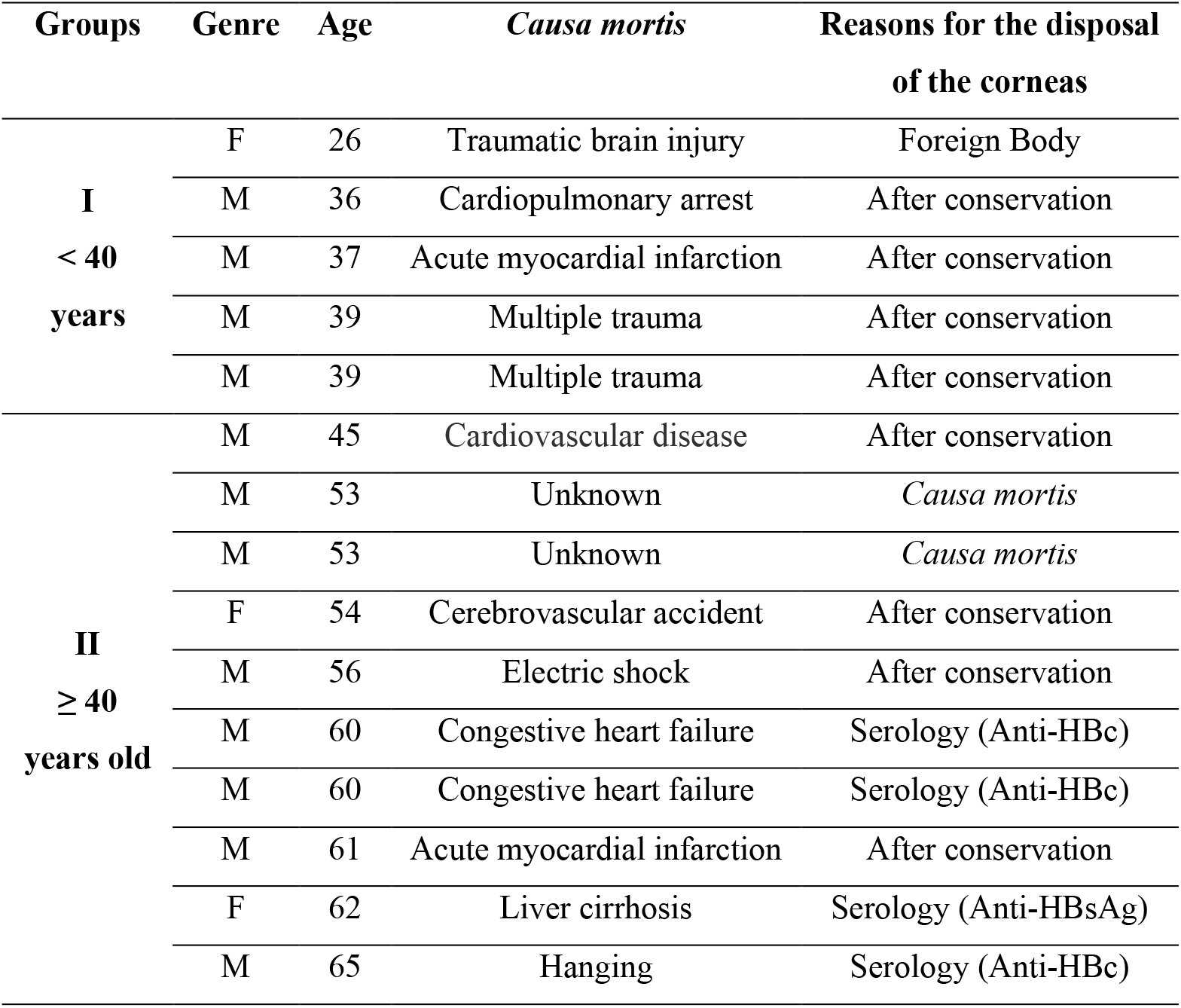
Epidemiological profile of individuals in groups I and II, *causa mortis* and reasons for the disposal of the corneas.

The ethical standards for scientific experimentation in humans established by the Helsinki Declaration (World Medical Association Recommendation 2000) and Resolution 196/96 of the Ministry of Health of Brazil (1996) were fully obeyed during the development of this study. We declare no conflicts of interest and ensure the confidentiality of the individuals participating in the study, which was approved by the Research Ethics Committee of the Faculty of Medicine of the University of Brasília, in September 9^th^ 2017 (CAAE n° 68498117.6.0000.5558).

### Sample processing

All the corneas were previously analyzed by Slit Lamp Biomicroscopy in order to assess thickness, opacity, presence of inflammatory infiltrates and vascularization of the corneal tissues, and by Specular Microscopy to evaluate the corneal endothelium.

After being collected in preservation solution, the corneas were sectioned in 3 parts and fixed in 2% glutaraldehyde solution and 2% paraformaldehyde in 0.1 M sodium cacodylate buffer, pH 7.2. After washing in cacodylate buffer and post-fixed in osmium tetroxide (1% in sodium cacodylate buffer) the specimens were dehydrated for 15 minutes in solutions with increasing concentrations of acetone (30%, 50%, 70%, 90 % and 3x 100%); after drying to the critical point with CO2 the histological specimens were metalized with gold to obtain the images by scanning electron microscopy (SEM) (JEOL 7001F, Tokyo, Japan).

### Morphological analyses

The analyzes were performed on the images obtained from the central and peripheral regions of each cornea using the GIMP 2.8 program (GNU Image Manipulation Program at http://gimp.org). The thickness of the corneal layers (anterior epithelium, Bowman’s layer, stroma, Descemet’s membrane and posterior epithelium) was taken at three regions equidistant and perpendicular to the Bowman’s layer, the integrity of the stromal lamellae was evaluated by the quantification of total interlamellar slits (5 x 5000 μm^2^) and the endothelial lesions were quantified in three areas of 500 μm^2^. The results were expressed in micrometers and compared between the two age groups (<40 years x ≥ 40 years).

### Statistical analysis

The results were evaluated using Bartlett’s test for equal variances and the Kolmogorov– Smirnov test for normal distribution before comparative analysis. Consider the unrelated samples, t-test or Mann-Whitney were used to compare two groups with normally or non-normally distribution. The Prism 5.0 software package (GraphPad, USA) was employed for statistical tests and graphical presentation of the data; differences with a two-tailed value of p < 0.05 were considered statistically significant.

## RESULTS

The results showed that for group I (<40 years) the thickness in the central region of the corneas is smaller than in the peripheral region (Test t; p = 0.039), whereas for Group II (≥ 40 years) there was no difference between these two regions (Mann-Whitney, p> 0.05). Also, there were no differences between the Group I and Group II in both peripheral (Mann-Whitney, p> 0.05) or central (Test t, p> 0.05) regions.

Layer analysis showed that in group I the Bowman, Descemet and endothelium layers were thicker at the periphery of the corneas than their respective central regions (Bowman / Mann-Whitney, p = 0.024; Descemet / test t, p = 0.013; Endothelium / Mann-Whitney, p = 0.012). For group II only the periphery layer of Bowman was thicker than its central region (Test t; p = 0.049) (Table 2).

**Table 2.** Thickness of the corneal layers obtained from two groups of individuals (< 40 years and ≥ 40 years). The analyses were made in the peripheral and central regions of each cornea and the medians are expressed in micrometers.

| Groups | Regions | Epithelium | Bowman | Stroma | Descemet | Endothelium | Total |
| --- | --- | --- | --- | --- | --- | --- | --- |
| **I**<br>**< 40 years** | Peripheral | 12.5 | 6.0* | 267.0 | 4.5*• | 3.5*• | 297.0* |
|  | Central | 7.3 | 4.0*• | 153.7 | 2.7* | 0.9* | 159.0* |
| **II**<br>**$\geq 40$ years** | Peripheral | 10.8 | 6.5* | 184.8 | 2.0• | 1.3• | 193.8• |
|  | Central | 3.0 | 5.0*• | 162.2 | 2.5 | 1.5 | 175.7 |

(\*) Peripheral region ≠ central region in the same group; (•) Group I ≠ Group II

Comparison between the two groups showed that the Bowman layer was thicker in the central region of the corneas of the group II than in group I (Test t, p = 0.032). In contrast, in group II the Descemet membrane and the endothelium were less thick than in group I (Descemet: test t, p = 0.047; Endothelium: test t, p = 0.015) (Table 2; Fig. 1).

**Figure 1.**
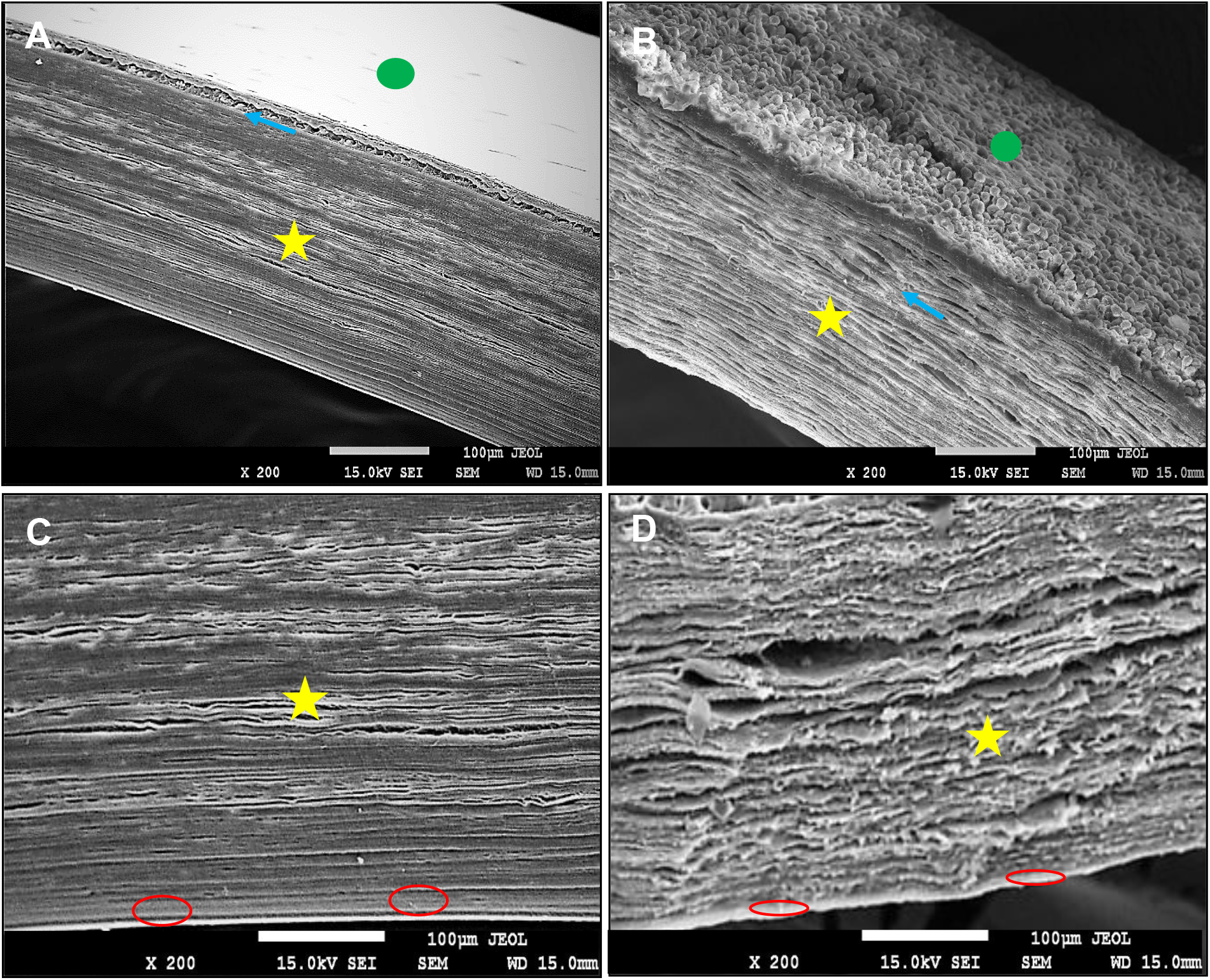
Eletromicrographys by SEM of human corneas obtained from individuals of the group I (A, C) and the group II (B, D) showing the anterior epithelium 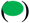, the Bowman layer 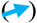, the stroma 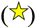 and the Descemet membrane 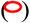. See the greater thickness of the Bowman in B and of the Descemet membrane in C.

The results showed a greater number of endothelial lesions in both regions of the corneas in group II (peripheral = 3.4 ± 2.3; central = 6.1 ± 3.5) than in group I (peripheral = 0.9 ± 1.2; central = 2.0 ± 2.5) (Test t, p <0.05). No difference was observed in the total number of interlamellar slits in both regions of the corneas in group I (peripheral = 5.4 ± 3.8, central = 7.2 ± 5.8) or in group II (peripheral = 6.9 ± 3.7, central = 7.2 ± 4.4) (T test, p> 0.05) (Fig. 2).

**Figure 2.**
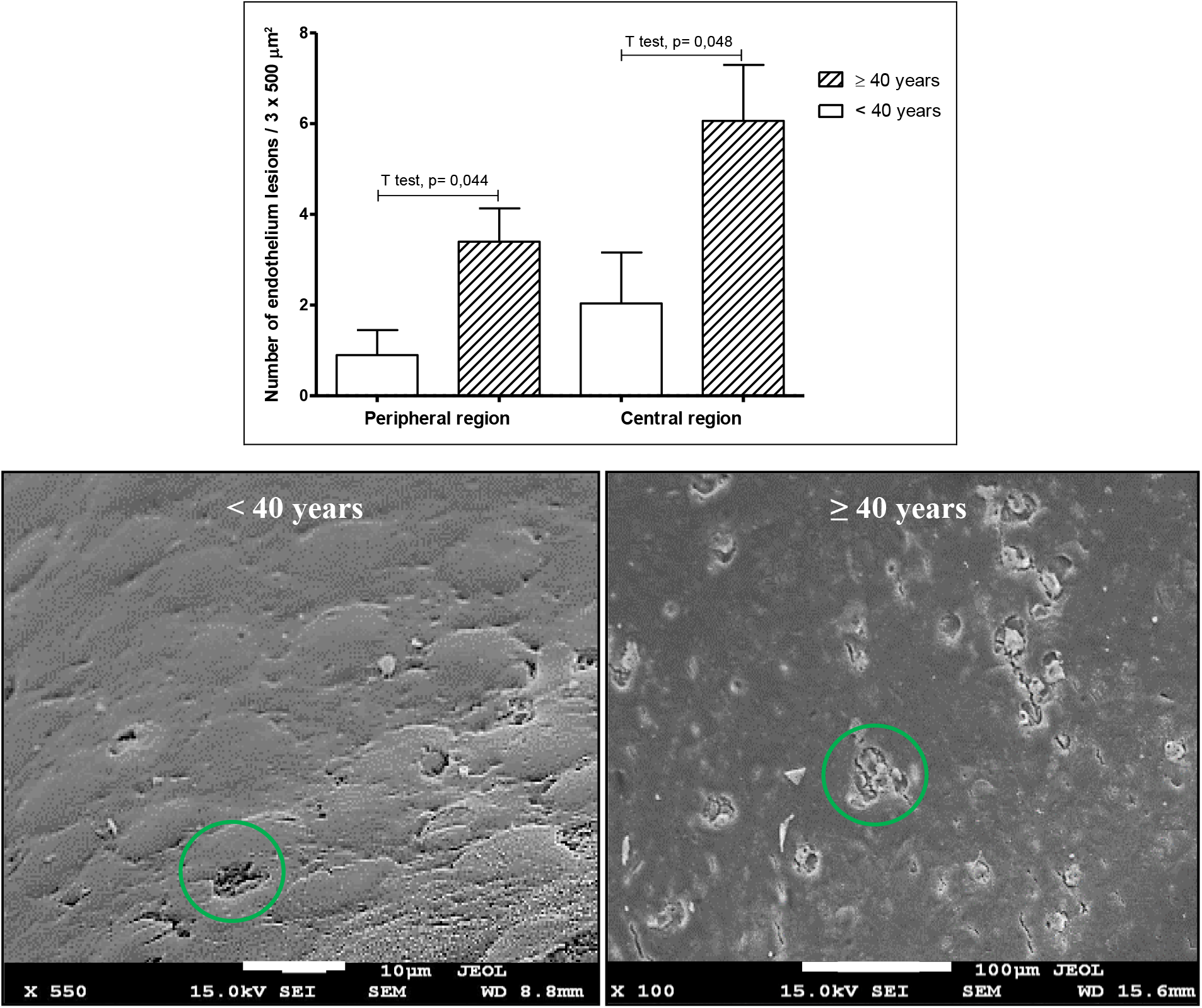
Total endothelial lesions quantified in an area of 3 x 500 μm^2^ in the corneas of the individuals of groups I (<40 years) and II (≥ 40 years). The results indicate a higher number of endothelial lesions in both peripheral and central regions of the cornea in group II (right figure – green circle), when compared to the group I (left figure). The mean and standard deviations are shown (above).

## DISCUSSION

Considering that the cornea is a fundamental structure in the vision and understanding how the aging process affects the shape and function of its layers can guide the therapeutic procedures, especially those that require surgical intervention. Our results showed that individuals aged 35 ± 5 years have thicker corneas in the peripheral region of the than in the central one. The finding that the periphery of the cornea is thicker in relation to its central region probably reflects its proximity to the limbic region than the region responsible for providing the stem cells that maintain and repair the epithelium and the corneal stroma (Sridhar, 2018).

Considering that aging reduces the pool of available stem cells for tissue renewal, it is expected that there will be a reduction in the corneal thickness also in the periphery of this organ and, in fact, our results showed that in the individuals with 57 ± 6 years there was a reduction in the thickness of the cornea when compared to the younger group; these results confirm the findings that corneal thickness decreases with aging (Lekskul et al., 2005; Rufer et al., 2007; Reinstein et al., 2008; Galgauskas et al., 2012). Despite all the corneal thicknesses were less than observed in vivo due to the dehydration process of the tissue, we highlight that it does not invalidate the study, since all the samples were prepared in the same way.

To identify which layers contributed to reduce corneal thickness in group II, their measurements were taken individually. Thus, our results showed that in the corneas of individuals aged 35 ± 5 years the Descemet membrane and the endothelium were thicker than in group II. In contrast, the Bowman layer was thicker in the central region of the corneas in individuals aged 57 ± 6 years when compared to younger individuals. The thickening of the Bowman layer identified in this study diverges from data reporting a reduction of 0.06 μm per year, reaching 1/3 of its initial thickness up to 80 years of age (Delmonte and Kim, 2011; Jacob and Naveen, 2016; Schlötzer-Schrehardt and Kruse, 2016). However, as individuals in group II are not actually elderly (51-63 years), tissue renewal in the anterior region of the cornea is expected to be functional, since fibroblasts (differentiated keratocytes) migrate from the anterior epithelium and maintain the structure of the Bowman layer and stroma.

Although not a statistical difference, a reduction in the thickness of the anterior epithelium (central region) was identified in group II that is suggestive of an initial process of cell loss. Under these conditions it is possible that there is thickening of the Bowman’s layer by the greater production of collagen by the fibroblasts as a compensation mechanism for the narrowing of the anterior epithelium. In contrast, the narrowing observed in the corneal layers in elderly individuals may be related to a reduction in the stem cell pool in the limbus region, thus reducing the contribution of these cells that are fundamental for the renewal of corneal tissues (Notara et al., 2013).

Of all the structural changes undergone by the cornea in aging, the most important and clinically relevant is endothelial cell loss estimated from 0.3% to 2.4% per year (Gipson, 2013; Arici et al., 2014; Elbaz et al., 2017; Sridhar, 2018). In this study, both endothelium and Descemet’s membrane in the corneas of the individuals of group II presented a reduction in the thickness, but in addition, the number of endothelial lesions exceeded those found in group I. The biological mechanisms of endothelial loss are lacking elucidation, but they may be related to low cell turnover rate, hormonal factors, environmental (ultraviolet radiation), chemical toxicity and oxidative stress (Sheng and Bullimore, 2007; Hatou, 2011; Jurkunas, 2018). In conditions of injury or even endothelial loss, the physiology of the cornea deteriorates and even the results of refractive surgeries are impaired (Dutt, 1994; Green, 1995; Galgauskas et al., 2014).

The corneas used in this study were collected from healthy individuals and potentially applicable to transplantation by corneal specular microscopy. However, after being analyzed by microscopy with large magnification, it was verified that the individuals aged 57 ± 6 years present endothelial lesions in greater numbers and a reduction in the thickness of the Descemet membrane, possibly related to mechanisms of compensation by the reduction in the stem cell pool of limbus that are responsible for the renewal of corneal tissues. These findings could compromise the success of refractive surgeries in individuals of group II and, despite the corneas being collected from healthy donors, they could also compromise the success of transplant. New investigations are suggested to identify possible cellular alterations such as the presence of apoptosis, vascularization and inflammatory infiltrate, as well as to elucidate how innervation, the anteroposterior distribution of the fibers of the elastic system in aging occurs. Our results point to an update of corneal uptake protocols for transplantation and may aid the interventions in refractive surgeries in individuals in different age.

## Data Availability

All data generated or analysed during this study are included in this published article.

## ACKNOWLEDGEMENT

We are grateful to Lucas Souza Kückelhaus, for reviewing the English language of the manuscript, the Laboratory of microscopy, Biology Institute, University of Brasilia, and to CAPES/Ministry of Education/Brazil for the financial support.

